# Ongoing evolution of SARS-CoV-2 drives escape from mRNA vaccine-induced humoral immunity

**DOI:** 10.1101/2024.03.05.24303815

**Authors:** Alex L. Roederer, Yi Cao, Kerri St. Denis, Maegan L. Sheehan, Chia Jung Li, Evan C. Lam, David J. Gregory, Mark C. Poznansky, A. John Iafrate, David H. Canaday, Stefan Gravenstein, Wilfredo F. Garcia-Beltran, Alejandro B. Balazs

## Abstract

Since the COVID-19 pandemic began in 2020, viral sequencing has documented 131 individual mutations in the viral spike protein across 48 named variants. To determine the ability of vaccine-mediated humoral immunity to keep pace with continued SARS-CoV-2 evolution, we assessed the neutralization potency of sera from 76 vaccine recipients collected after 2 to 6 immunizations against a comprehensive panel of mutations observed during the pandemic. Remarkably, while many individual mutations that emerged between 2020 and 2022 exhibit escape from sera following primary vaccination, few escape boosted sera. However, progressive loss of neutralization was observed across newer variants, irrespective of vaccine doses. Importantly, an updated XBB.1.5 booster significantly increased titers against newer variants but not JN.1. These findings demonstrate that seasonal boosters improve titers against contemporaneous strains, but novel variants continue to evade updated mRNA vaccines, demonstrating the need for novel approaches to adequately control SARS-CoV-2 transmission.

## INTRODUCTION

Since it was first described in late 2019, SARS-CoV-2 has undergone continuous evolution. The virus originated in Wuhan, China, and was declared a global pandemic on March 11, 2020 by the WHO^1,2^. Since then, it has infected over 700 million people and caused nearly 7 million deaths^2,3^. The first single mutation to become fixed in the population, D614G, emerged in February of 2020 and was shown to be more infectious and stable than the original sequence^4,5^. However, the first variant to be named, B.1.1.7 or Alpha, harbored multiple mutations and became the dominant strain in the UK, exhibiting over 50% greater infectivity than wild-type (WT)^6^. As the pandemic entered its second year, additional variants emerged carrying mutations in the receptor binding domain (RBD) that enhanced transmissibility^7,8^ through improved ACE2 binding and exhibited viral escape from convalescent sera^9–11^. In the subsequent two years, the number of variants that have been documented has increased significantly, with new mutations (predominantly in the RBD) that enable escape from vaccine-induced neutralizing antibodies and enhance transmissibility^10,12–15^.

An unprecedented effort to develop effective countermeasures resulted in multiple vaccines being approved by the FDA in the United States, all of which were based upon the WT SARS-CoV-2 spike protein containing stabilizing mutations that were previously identified for RSV and MERS^16,17^. Numerous vaccine technologies were employed, including mRNA containing lipid nanoparticles, recombinant proteins, or adenovirus vectored vaccines^18–21^, however mRNA vaccines were the most widely deployed in the United States. Clinical trials of these vaccines demonstrated remarkable efficacy to reduce COVID-19 infections, hospitalizations, and deaths^18–21^.

However, the emergence of the Omicron variant and declining antibody titers over time led to the recommendation of a third booster dose of mRNA vaccine by the FDA, yielding neutralizing, cross-reactive antibodies against Omicron^13,22^. The emergence of new variants such as BA.5 in 2022, necessitated the reformulation of vaccines^23^, spurring the transition from monovalent to bivalent vaccines incorporating both WT and BA.5 spikes^24^. Despite the efficacy of the original vaccines in preventing severe disease and death, their effectiveness waned against heavily mutated, highly transmissible variants such as BQ.1.1, XBB, BA.2.86 and their sub-variants^15,25–27^. These highly evolved strains carried 34-58 total mutations within the spike protein, with 28 of these mutations in the RBD in the most recent circulating variant JN.1. Recently, an FDA advisory committee recommended deployment of an updated monovalent mRNA vaccine encoding the XBB.1.5 spike^28^. However, a variant named BA.2.86 has arisen recently that harbors 57 total spike mutations, including 20 that have not been reported previously, raising concerns that derivatives of this strain could escape the XBB.1.5 booster. BA.2.86 and its derivatives including JN.1 now make up over 90% of new COVID sequences being submitted to GISAID^29,30^.

To understand the impact of mutations which have arisen across the shifting vaccination landscape, we conducted a comprehensive analysis of individual mutations found in variants of concern or interest which emerged during the pandemic. Using a previously validated high throughput neutralization assay^10,13,31^, we measured the neutralization of pseudoviruses representing each individual mutation across study participants who had received either the primary vaccine series (two shots) or the booster series (three shots). Furthermore, we evaluated the neutralization activity of sera from boosted individuals against all individual mutations, as well as strains harboring combinations of mutations through JN.1, totaling 220 different pseudoviruses. Interestingly, we find that a handful of individual mutations arising after Omicron enable significant escape from boosted serum. Remarkably, the most recent variants, including BQ.1.1, XBB, and XBB.1.5, exhibit escape from boosted sera that is comparable to SARS-CoV-1 and the related pre-emergent WIV1 strain. However, sera from individuals receiving bivalent boosters exhibited significant neutralizing activity against more recent strains. Despite this, more recent variants, such as EG.5.1 and HK.3, exhibit substantial escape from bivalent boosted sera, suggesting that SARS-CoV-2 remained ahead of efforts to update vaccines. We find that the latest XBB.1.5 booster enhances neutralizing activity against many variants that escaped bivalently boosted sera, however the latest JN.1 variant remains significantly resistant to neutralization across vaccinations. Together, these results highlight the importance of continued surveillance of SARS-CoV-2 sequence evolution and support the updating of existing vaccines. At the same time, our results also highlight the need for novel approaches capable of inducing broadly neutralizing humoral immunity to counter the continued evolution of SARS-CoV-2 variants.

## RESULTS

### SARS-CoV-2 variants of concern have accumulated spike mutations focused within RBD

SARS-CoV-2 has resulted in over 700 million infections and nearly 7 million deaths since it first emerged in 2020^2,3^. The virus, which originated in Wuhan China, circulated for nearly a year before the first variant of concern (Alpha) emerged (**Figure 1A, Table S1**), however in subsequent years, numerous variants of concern and interest have been described (**Figure 1B, Table S1**). One of the most globally dominant variants, Delta (B.1.617.2), was ascendant in June 2021 and harbored 8 mutations, including 2 in the RBD^6^. However, the Omicron variant (BA.1), containing 32 spike mutations, rapidly rose to prominence after it first appeared in November 2021. Variants derived from BA.1 continued to emerge resulting in the BA.5 strain harboring two additional mutations within the RBD. Most recently, JN.1 has become the dominant strain globally^30^, harboring 58 total spike mutations with 27 of these in the RBD^29^. Impressively, the SARS-CoV-2 pandemic has now resulted in variants that are further evolved from the original Wuhan strain than other distinct coronaviruses such as RaTG13^32,33^ (**Figure S1A**) which harbors only 26 amino acid differences from the Wuhan strain. Over the course of the pandemic, 131 differences in viral spike have arisen across variants of concern/interest, including several which occurred at the same position in the spike protein (**Figure S1B**).

**Figure 1:**
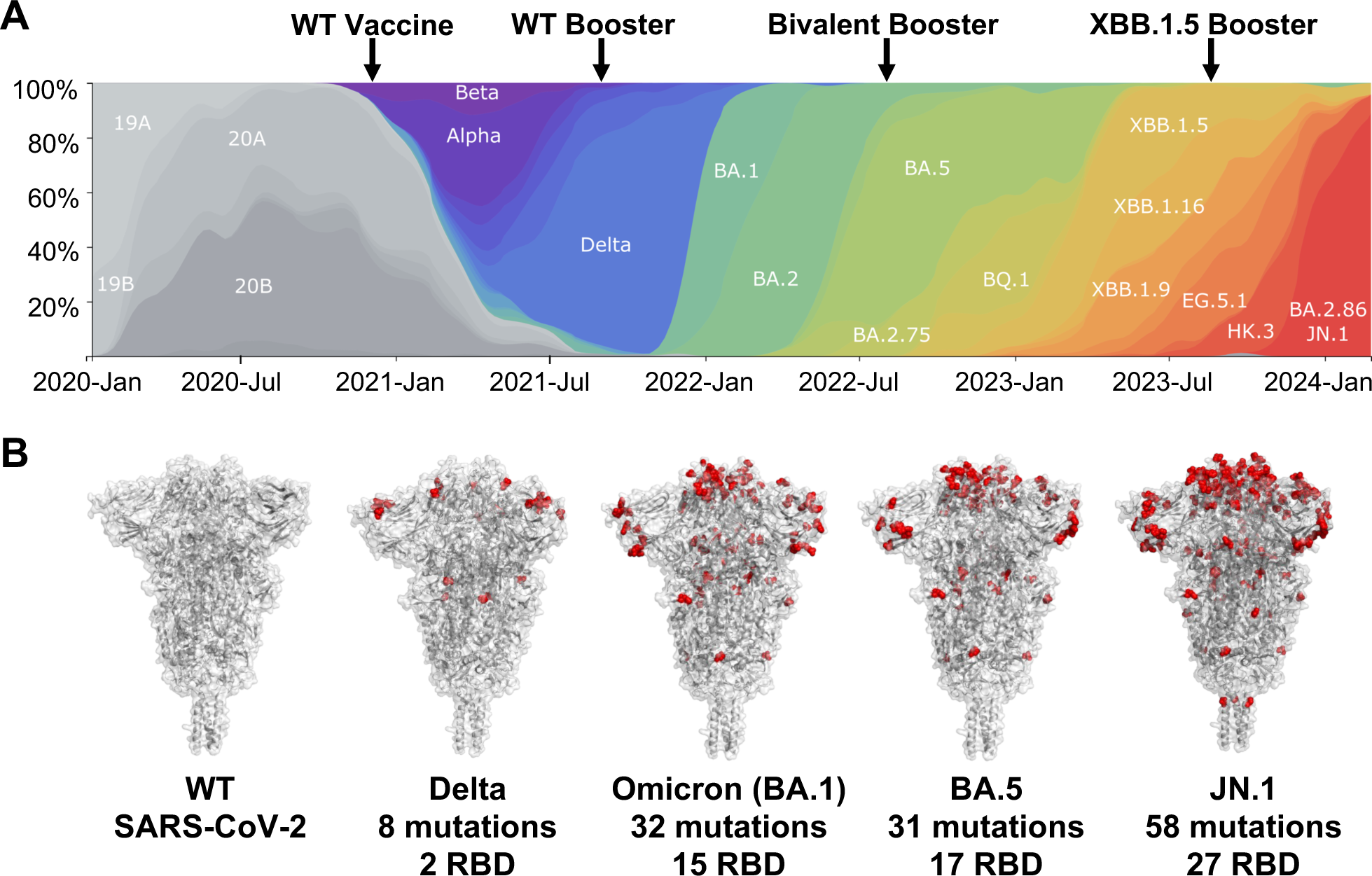
SARS-CoV-2 variants of concern harbor spike protein mutations that are enriched in the RBD. (A) Scaled stacked area chart of variants emerging during the SARS-CoV-2 pandemic. Arrows denote when each vaccine was made available. Adapted from nextstrain.org. Data as of February 28^th^, 2024. (B) Crystal structures of pre-fusion stabilized SARS-CoV-2 spike trimer (PDB ID 6XR8) highlighting mutations found in each strain mapped onto their locations on the wild-type spike trimer. From left to right WT, Delta (B.1.617.2), Omicron (BA.1), BA.5, and JN.1 strains. Mutations highlighted for each strain are from corresponding table S1.

### Neutralization assays with primary vaccine sera reveals regions of vulnerability within the natural mutation landscape

To investigate the effect of all individual spike variations on vaccine induced humoral immunity we studied a cohort of 20 serum donors who had received the primary vaccine series (two shots of Pfizer (BNT162b)). The cohort had a median age of 33 and was 50% female. Sera from this group was subjected to pseudovirus-based neutralization assays that we and others have previously validated^10,13,31,34–38^. In brief, pseudotyped lentiviral particles bearing a given SARS-CoV-2 spike protein and encoding a luciferase-expressing transgene were produced and mixed with serially diluted donor serum before ACE2-expressing target cells were added (**Figure 2A**). Assays were performed using pseudoviruses representing 69 differences present across 24 named variants of concern/interest up-to and including Delta. Variants with multiple consensus sequences such as Beta (V1-V6) are denoted as separate variants (**Figure S2**). Donor serum exhibited high neutralizing activity against the Wuhan pseudovirus, with a geometric mean pseudovirus 50% neutralization titer (pNT_50_) of 2962 (**Figure 2B**). However, individual changes in either the N-terminal domain (NTD), RBD or portions of the S1 region exhibited significant escape from neutralization. Within the RBD in particular, K417N, K417T, L452R, and Y453F had highly significant reductions (P<0.0001) in neutralization titers with geometric means of between 300-500. Other mutations in the NTD and S1 subunit significantly reduced pNT_50_ including P26S, T29I, Q52R, D80A, T572I, Q677H, and P681R while those found within the S2 domain had little effect on neutralization titers except V1176F and M1229I. To explore patterns of individual vaccine responses, two-way hierarchical clustering was performed across all mutations tested for each individual donor (**Figure 2C**). This analysis revealed that individual sera had remarkably similar loss of neutralization activity across a subset of mutations within the RBD, NTD, and S1 with more than half of all mutations being harder to neutralize than WT. Interestingly, two separate clusters of donors were observed in this cohort, with the bottom cluster having a broader overall response to individual mutations arising through the Delta strain. However, we found no correlation across time, age or sex to account for vaccination outcomes (not shown). These results suggest that humoral responses in most individuals were focused on particular epitopes within the NTD, RBD and S1, following the primary vaccination regimen with approximately half of these individuals generating broader responses.

**Figure 2:**
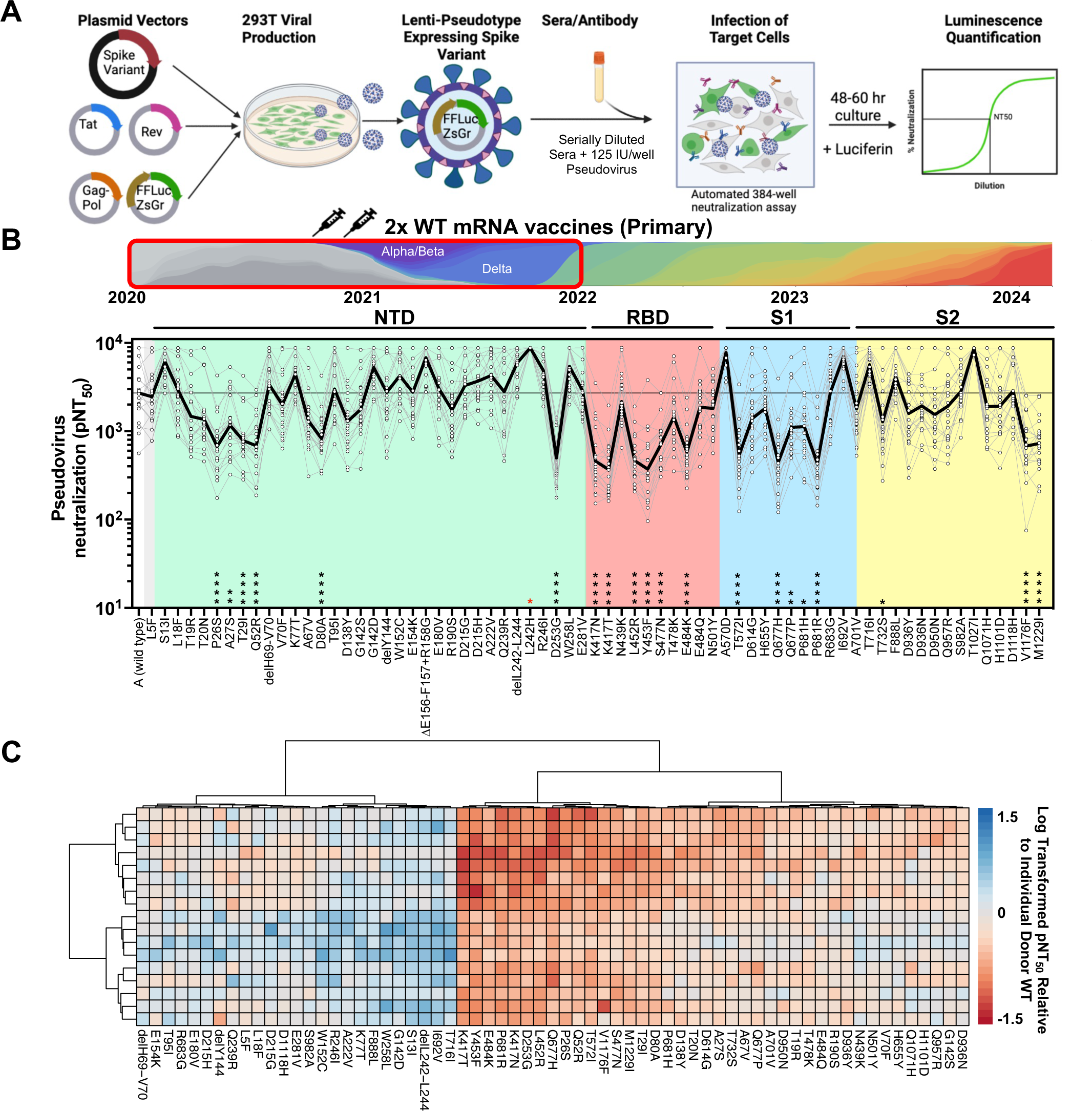
Neutralization assays with primary vaccine sera revealed regions of vulnerability within the natural mutation landscape. (A) Schematic of high throughput neutralization assay used in these studies. (B) (Top) Schematic illustrating the time period from which variants were selected and the vaccinee sera to be tested below. (Bottom) Titers that achieve 50% of pseudovirus neutralization (pNT_50_) are plotted for all individual mutants found in SARS-CoV-2 through Delta (1.617.2) for 24 COVID naive donors who received the primary vaccination regimen (2 doses of Moderna or Pfizer). The solid black line represents geometric mean pNT_50_ for reference. The following abbreviations are used NTD = N-Terminal Domain, RBD = Receptor Binding Domain, S1 = S1 Subunit, S2 = S2 subunit. pNT_50_ of each spike mutant was compared to pNT_50_ of wild type using a One Way ANOVA with Friedman’s test and Dunn’s multiple comparisons test. (* = P<0.0332, ** = P<0.0021, ***= P<0.0002, ****= P<0.0001, red = significantly higher than WT). (C) Two-way hierarchical clustering of pNT_50_ values of sera obtained from vaccinated donors (rows) across each variant (columns) relative to individual donor WT titer. pNT_50_ values are plotted in a heat map colored according to neutralizing activity relative to WT. Donors with WT neutralizing titers at the maximum of our assay were excluded. Mutations where a significant majority of donor titers reached the limit of detection were also excluded. Clustering was performed using pheatmap package v1.0.12 in R-Studio.

### Combinations of individual mutations contribute to neutralization resistance

Given that variants of concern/interest harbor multiple mutations, we assessed the impact of each individual mutation in the spike of the Beta variant. Of note, no individual mutation could account for the differences in pNT_50_ between WT and Beta variant spike, but K417N and E484K were both highly neutralization resistant (**Figure 3A, left**). We previously showed that the RBD region of Beta was largely responsible for escape from neutralization^10^. To understand how multiple RBD mutations might coordinate to escape humoral immunity, primary vaccine sera was tested against double and triple RBD mutants (**Figure 3A, right**). Neither single mutation came close to recapitulating the neutralization resistance of the parent spike, but we did observe an additive effect for the three individual RBD mutations found in the Beta variant. Combinations of any two mutations yielded greater resistance to neutralization than single mutations, but the triple mutant exhibited nearly as much escape as the full Beta spike. Despite the neutralization resistance of Beta variant pseudovirus, genomic surveillance suggests that the Delta variant rapidly overtook all other circulating SARS-CoV-2 variants over a period of eight months^30,39,40^. Primary vaccine sera was also assessed against each of the individual Delta variant mutations, with L452R and P681R exhibiting significant reduction of pNT_50_ (**Figure 3B, left**). In contrast to Beta, combining RBD mutations in Delta resulted in no additional reduction in pNT_50_, suggesting that mutations outside of RBD, namely P681R, appear to coordinate with L452R to drive greater escape (**Figure 3B, right**).

**Figure 3:**
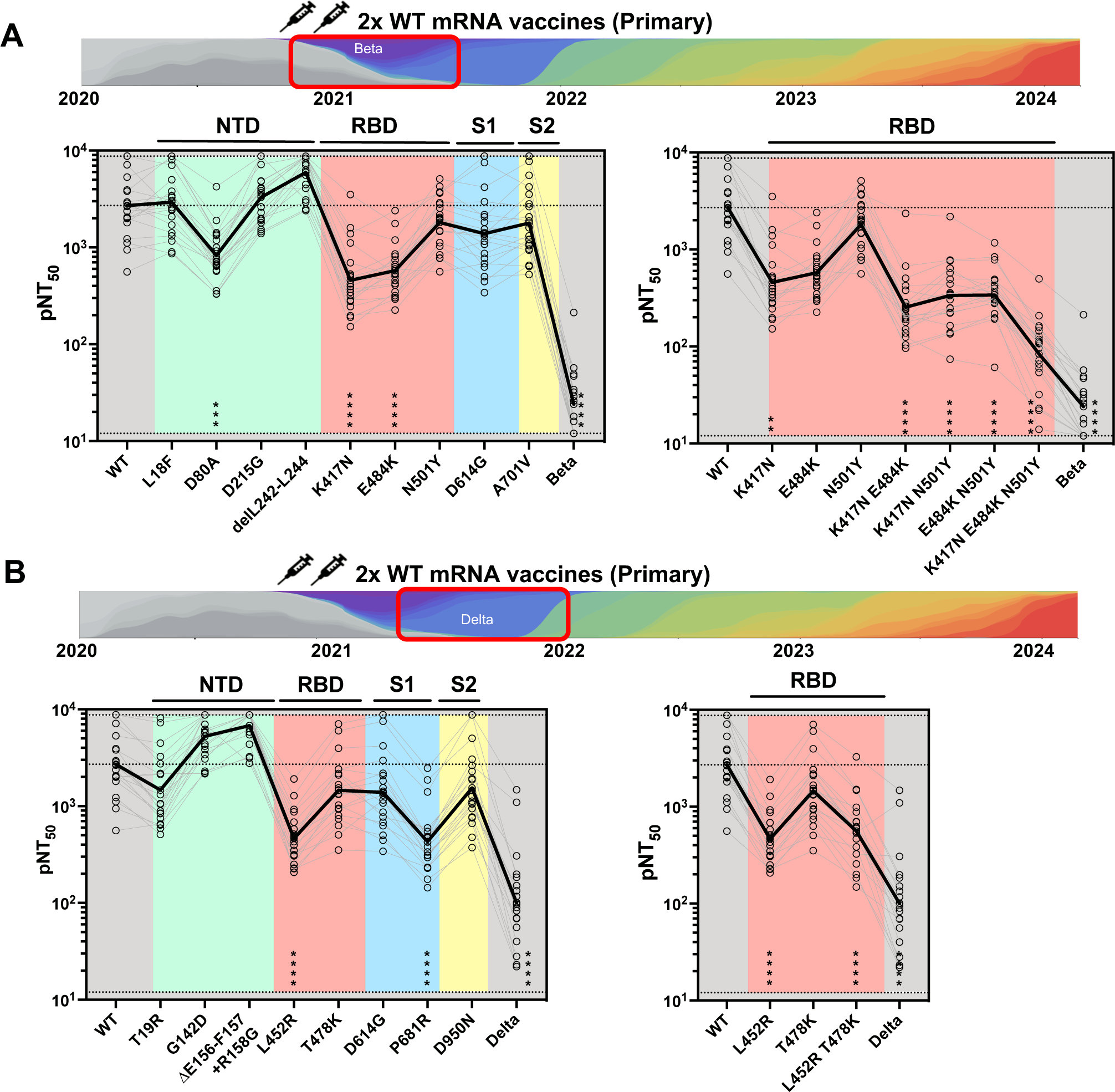
Combinations of individual mutations yield neutralization resistance. (A-B) Primary series vaccinee sera was tested via pseudovirus-based neutralization assays against each individual mutation or combination of mutations found in the (A) (Left) Beta strain or (B) (Left) Delta strain. Each variant was compared to the wild type using a One-way ANOVA with Friedman’s Test and Dunn’s multiple comparisons test. (* = P<0.0332, ** = P<0.0021, ***= P<0.0002, ****= P<0.0001). Primary series vaccinee sera was tested against combinations of (A) (Right) Beta or (B) (Right) Delta RBD mutant pseudoviruses and compared to the complete set of mutations for each strain. Each group was compared to wild type using a One-way ANOVA with Friedman’s Test and Dunn’s multiple comparisons test. (* = P<0.0332, ** = P<0.0021, ***= P<0.0002, ****= P<0.0001).

Similar analyses were performed for other designated variants of concern (VOC) and their individual spike mutations including the Alpha (**Figure S3A**), Gamma (**Figure S3B**), and Epsilon variants (**Figure S3C**). Among these, all variant spikes exhibited reduced neutralization by sera from primary vaccination, likely from individual mutations within S1 for Alpha, RBD and NTD for Gamma and the RBD for Epsilon. Taken together, our results indicate that multiple mutations often coordinated to reduce neutralizing activity against variants of concern following primary vaccination.

### mRNA booster vaccination significantly enhances neutralization of SARS-CoV-2 mutants

During a significant spike in COVID-19 cases in July 2021 stemming from the Delta variant, a third mRNA booster shot consisting of the same Wuhan spike formulation as the primary series was introduced in the United States^41^ (**Figure 1A**). To measure the impact of a third mRNA administration on humoral immunity, 20-22 samples from primary vaccinee or mRNA boosted donors were matched for age and subjected to neutralization assays across the same set of individual mutations reported through October 2021 (up-to the Delta variant) (**Figure S4A, S5**). The vast majority of mutants tested were more potently neutralized relative to the WT titer by serum from boosted donors than those who had only received the primary series (**Figure 4A**). Two-way hierarchical clustering was performed across all mutations tested for each individual donor (**Figure 4B**). Boosting greatly enhanced the breadth of neutralizing activity relative to WT titer for all except one donor (**Figure 4B**). Across nearly all variants tested, there was improved neutralization potency relative to wildtype, suggesting an overall broadening of the humoral response (**Figure 4C**). Interestingly, some mutants, including D80A, K417N, T572I, and Q677H, exhibited disproportionately greater neutralization, suggesting that these epitopes were preferentially targeted by booster vaccination (**Figure 4C**). Comparison between the two groups found that donors who received the booster vaccine had significantly increased neutralization activity overall (**Figure 4D**). Comparing neutralization activity across mutations positioned within different spike regions showed significantly increased activity in boosted samples across spike regions, including NTD (**Figure S4B**), S1 (**Figure S4C**), RBD (**Figure S4D**) and S2 (**Figure S4E**).

**Figure 4:**
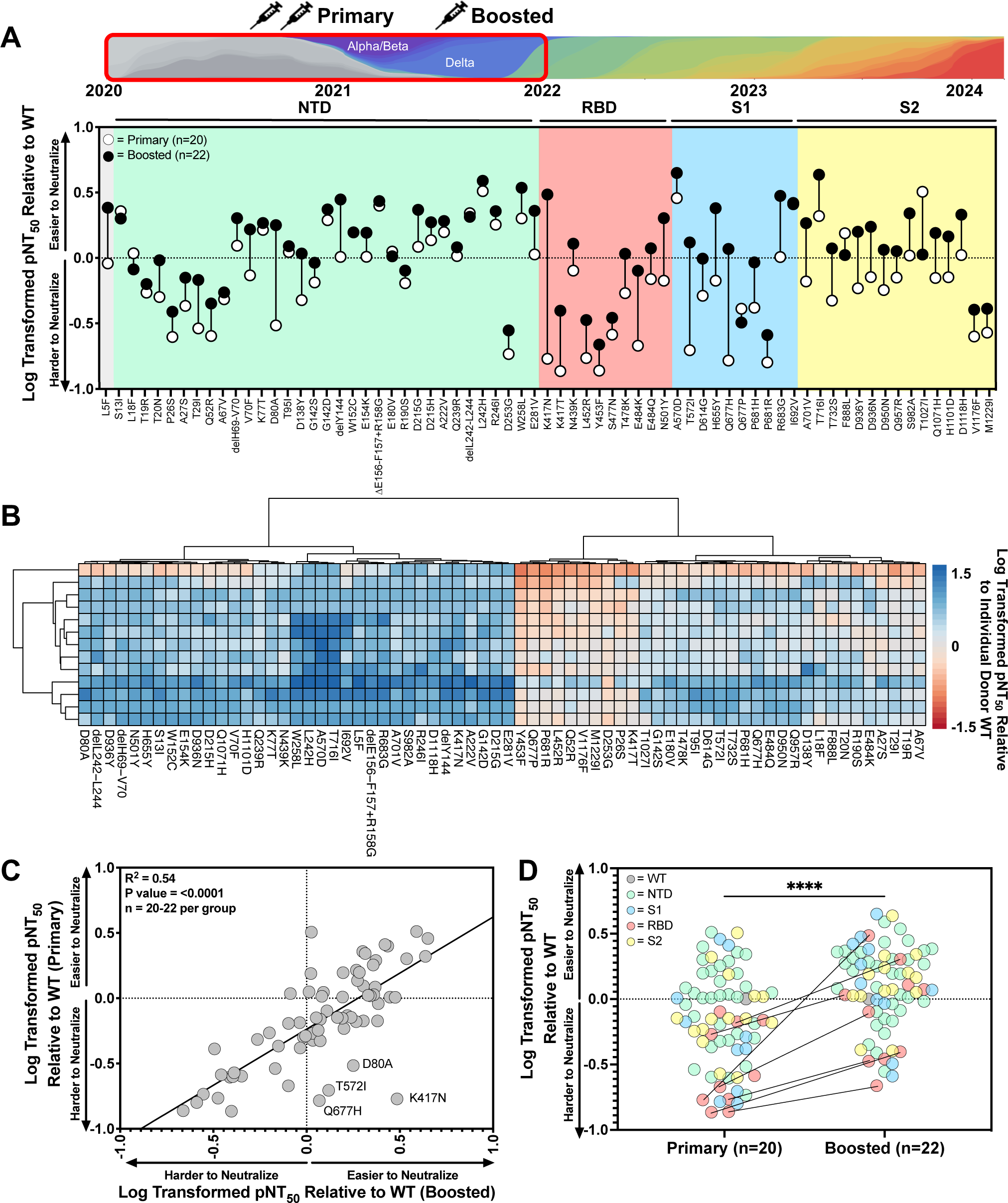
Individual Omicron variants harbor novel sites of vulnerability, while recent strains exhibit significant escape from boosted sera. (A) (Top) Schematic illustrating the time period from which variants were selected and the vaccinee sera to be tested below. (Bottom) Primary or Boosted pNT_50_ for donor across each variant was normalized to geometric mean titer across all donors for WT and log transformed. Length of each vertical line illustrates pNT_50_ change. (B) Two-way hierarchical clustering of pNT_50_ values of sera obtained from boosted donors (rows) across each variant (columns) relative to individual donor WT titer. pNT_50_ values are plotted in a heat map colored according to neutralizing activity relative to WT. Donors with WT neutralizing titers at the maximum of our assay were excluded. Mutations where a significant majority of donor titers reached the limit of detection were also excluded. Clustering was performed using pheatmap package v1.0.12 in R-Studio. (C) Log-transformed pNT_50_ geometric mean values for each variant relative to WT for primary series were plotted against matching boosted values. Linear regression (R^2^ = 0.5109; Slope = - 0.098; P<0.001) (D) Log-transformed pNT_50_ geometric mean values for each variant relative to WT colored according to their location in the spike protein comparing primary to boosted vaccinee sera. Statistics represent Wilcoxon two-tailed paired, non-parametric t-test with **** = P<0.0001. Selected RBD mutations from the delta and beta strains connected with lines showing their respective values for primary or boosted donor serum.

### Recent variants reveal progressively more escaped mutations from boosted sera

In November of 2021, a SARS-CoV-2 variant harboring an unusually large number of mutations was first identified in South Africa^42^. This novel variant of concern, designated Omicron (BA.1), contained 59 mutations in its genome, 32 of which were in the spike protein. The heavily mutated spike contained 15 RBD mutations, raising the possibility that it had escaped from humoral immunity. A number of heavily mutated, highly transmissible variants have descended from BA.1 since this time^25,43,44^. Additional neutralization assays were performed to assess whether boosted sera exhibited improved activity against more recent mutations (**Figure 5A, Figure S6**). While sera from individuals receiving a third mRNA booster vaccine was able to neutralize most mutants, spike mutations V445P, N460K and F486P exhibited 8.6-, 6.5-, and 7.7-fold decrease in pNT_50_ respectively (**Figure 5A**). Across the full set of mutations since the beginning of the pandemic, a number of individual mutants in the RBD escaped from mRNA-boosted serum samples (**Figure S6, Figure S7A**). Compared to mutations that had emerged in earlier variants including Delta, those emerging in Omicron and subsequent variants demonstrated significantly less neutralization by boosted sera (P = 0.0021) (**Figure 5B**). Mapping these mutations onto the structure of the SARS-CoV-2 spike showed that mutations appearing after Delta were largely concentrated at the apex of the spike and specific residues within the NTD (**Figure 5C**). Mapping the neutralization resistance of all 131 mutations onto the spike protein crystal structure revealed that the most escaped mutants appeared at the apex of the spike and on the outer edges of the NTD (**Figure S7B**). Neutralization values against RBD mutants over time revealed a significant reduction in pNT_50_ relative to WT (P = 0.02), suggesting that more recent mutations drove escape from vaccine responses (**Figure 5D**). Analysis of boosted neutralizing titers against mutations over time showed an overall trend towards greater escape (R^2^ = 0.08, P = 0.0006), but relatively few novel mutations have been described in variants of concern/interest between 2022 and 2023 (**Figure S7C**). Neutralization of more recent strains showed a 5-fold decrease in pNT_50_ value beginning with BA.1, further decreasing to 104-fold for the most escaped among them, BQ.1.1.22 (**Figure 5E**). Two-way hierarchical clustering was performed across all mutations through the XBB.1.16 variant for boosted neutralizing titers (**Figure S7D**). Mutations that clustered together that were hardest to neutralize are all present in variants that descended from XBB. Donors separated into two clusters, one with a broader overall response and another cluster with notable weaknesses against mutations present in XBB variants.

**Figure 5:**
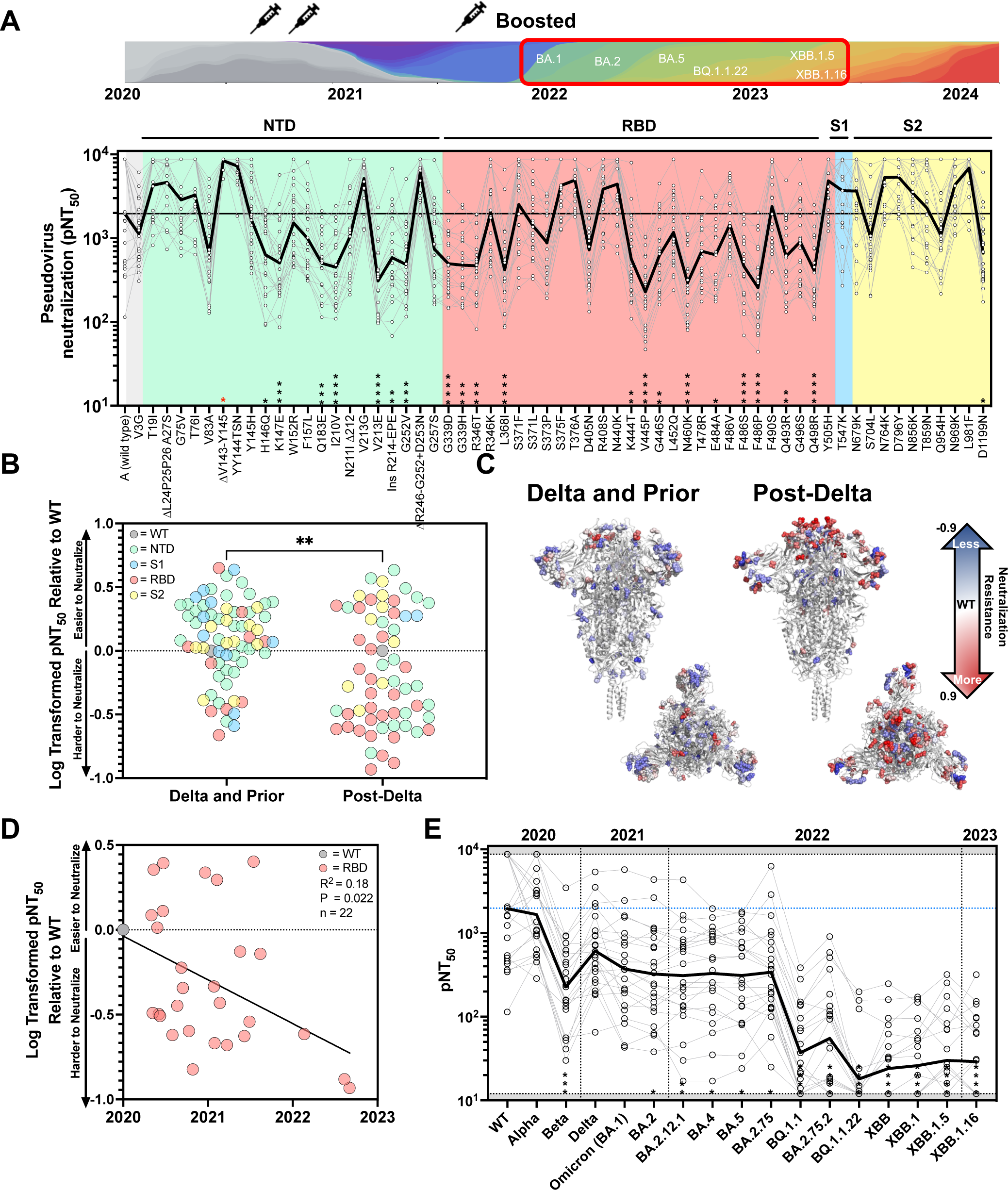
Individual Omicron variants reveal new sites of resistance that are observed in more recent strains to significantly escape from boosted sera. (A) (Top) Schematic illustrating the time period from which variants were selected and the vaccinee sera to be tested below. (Bottom) pNT_50_ are plotted for all individual spike mutations occurring after Delta (1.617.2) through XBB.1.16 for 24 COVID naive donors that received primary vaccination (2 doses of Moderna or Pfizer) and a booster shot. The solid black line indicates the geometric mean of the titers against each pseudovirus. The following abbreviations are used NTD = N-Terminal Domain, RBD = Receptor Binding Domain, S1 = S1 Subunit, S2 = S2 subunit. Each pseudovirus was compared to SARS-CoV-2 wild type using a One Way ANOVA with Kruskal Wallis Test and Dunn’s multiple comparisons test (* = P<0.0332, ** = P<0.0021, ***= P<0.0002, ****= P<0.0001, red = significantly higher than WT). (A) (B) Log-transformed geometric mean pNT_50_ values normalized to WT for mutations that appeared in strains between WT and Delta as compared to those appearing post-Delta to XBB.1.16. Statistics represent Mann-Whitney two-tailed unpaired, non-parametric t-test with ** = P < 0.0021. In this case P = 0.0010. (B) The corresponding spike crystal structures (PDB 6xr8) from (B) with mutations colored according to neutralization resistance to boosted sera. (Left) Highlighting mutations between WT and Delta and (Right) highlighting mutations post-Delta to XBB.1.16. Log transformed values for each mutation in the spike are plotted on a three-color scale with a spectrum of -0.9 to 0.9. (C) Log transformed pNT_50_ geometric mean values relative to WT for boosted donor sera against all RBD variants in strains that appeared after Delta plotted against the date of first submission to GISAID. (D) pNT_50_ values for boosted donor sera against variants that appeared post-Delta to XBB.1.16, with Alpha, Beta, and Delta for reference. Strains are plotted in the order that they first appeared. The solid black line indicates the geometric mean of the titers against each pseudovirus. Each pseudovirus was compared to SARS-CoV-2 WT using One Way ANOVA with Kruskal Wallis Test and Dunn’s multiple comparisons test (* = P<0.0332, ** = P<0.0021, ***= P<0.0002, ****= P<0.0001).

### Bivalent and XBB.1.5 boosters markedly improve neutralization against recent high-Infectivity variants

During a surge in cases, largely attributed to the BA.5 variant, bivalent mRNA vaccines encoding both WT and BA.5 spike sequences were approved for use in the United States in August of 2022^23,45^. With the recombination of spikes producing XBB and sub variants, and the waning immunity of bivalent vaccination against these variants^46,47^, a newly reformulated vaccine encoding only the XBB.1.5 spike sequence was recommended for use in September 2023^48,49^. We obtained longitudinal samples from 29 nursing home residents (median of 72 years old) and 7 health care workers (median age 60) who received either three or four WT mRNA vaccines followed by a bivalent booster and an XBB.1.5 booster. To measure the impact of each vaccine administration, longitudinal serum samples were tested against variants previously found to escape boosted sera, or which were dominant variants for more than a month (**Figure 6A**). While there was a clear trend of increased neutralizing activity with each successive vaccination, only modest differences were observed between the 3rd and 4th WT mRNA boost (**Figure 6B**). Bivalent vaccines improved pseudovirus neutralization titers after BA.5, though the activity against these variants was still significantly lower than WT (**Figure S8A**). In contrast, only the most recent dominant variant, JN.1, and the original Beta variant showed significant escape from the XBB.1.5 boosted samples (**Figure S8B**). Overall, we observed increasing neutralization titers against all pseudoviruses tested relative to WT in the same cohort across each successive vaccination from WT booster to bivalent booster, and finally to XBB.1.5 booster (**Figure 6C**). We also observed a significant decline in neutralization activity across variants that arose across time for WT and Bivalent boosters but no significant decline across variants following XBB.1.5 booster vaccination (**Figure 6D**). Two-way hierarchical clustering was performed on matched donors at the boosted time point to understand the effect of additional boosters against new vaccinations (**Figure 6E**). Donors separated into two clusters after receiving a 3rd WT booster; one with broader neutralizing activity and another with lower activity against BQ.1.1.22 and later strains. However, the distinction between these two clusters was largely attenuated with each successive administration of Bivalent and XBB1.5 boosters.

**Figure 6:**
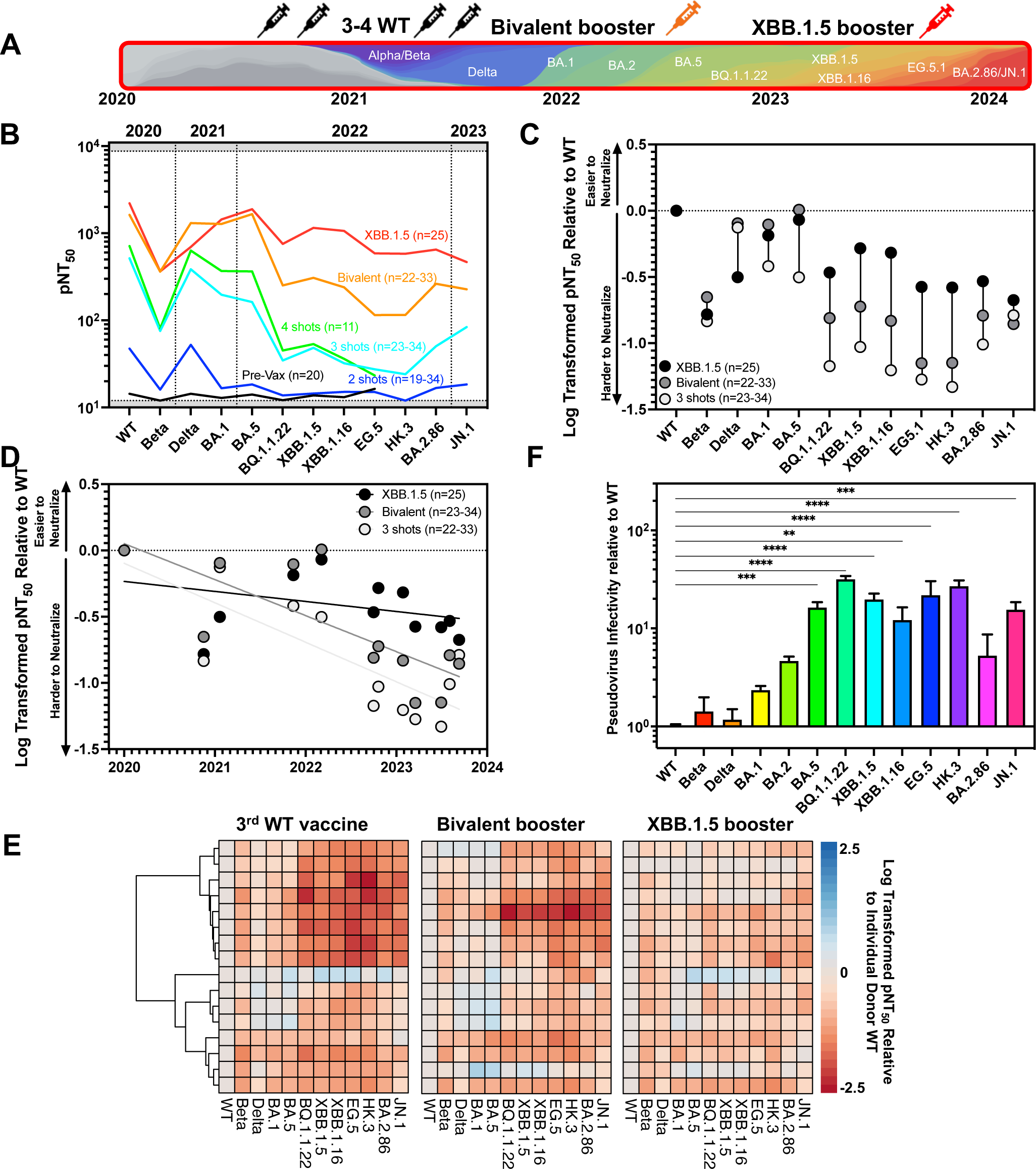
Bivalent and XBB.1.5 boosters improve neutralization against highly infectious variants. (A) Schematic illustrating the variants and vaccinee sera to be tested below. (B) Geometric mean pNT_50_ values of longitudinal serum samples over the course of CDC-recommended vaccination schedule against each variant. Strains are plotted in the order in time that they first appear. (C) pNT_50_ of individual serum samples were normalized to geometric mean titer across all donors for WT and log transformed. Longitudinal samples from donors after WT, Bivalent, or XBB.1.5 boosters are plotted. Length of each vertical line illustrates pNT_50_ change. (D) Log transformed pNT_50_ geometric mean values relative to WT for donor sera against variants of concern through JN.1 plotted against the date of first submission to GISAID. Solid lines represent linear regressions for WT Booster (R^2^=0.63, slope=-0.298, P=0.002), Bivalent Booster (R^2^=0.57, slope=-0.27, P=0.0044) and XBB.1.5 booster titers (R2=0.14, slope =-0.076, P =0.23). (E) One-way hierarchical clustering of pNT_50_ values of sera obtained from boosted donors (rows) across each variant of concern (columns) relative to individual donor WT titer. Heatmaps maintaining clustering order of rows across Bivalent and XBB.1.5 boosters are plotted to the right. pNT_50_ values are plotted in heat maps colored according to neutralizing activity relative to WT. Clustering was performed using pheatmap package v1.0.12 in R-Studio. (F) Pseudovirus infectivity (defined as infectious units per genome copy) was normalized to WT infectivity for each variant spike across three technical replicates (n=3). Pseudovirus infectivity relative to WT was measured for major SARS-CoV-2 variants by calculating fold change in slope of linear regressions fit across known dilutions. Bars and error bars depict mean and standard error of the mean. Each pseudovirus was compared to SARS-CoV-2 wild type using a One Way ANOVA with Holm-Sidak correction for multiple comparisons. (* = P<0.033, ** = P<0.0021, ***= P<0.0002, ****=P<0.0001).

Given the vast number of mutations in new variants, we wanted to assess their effect on the efficiency of viral entry. To determine the viral entry efficiency of variant spikes, we compared the rate of pseudovirus transduction over a defined range of concentrations (**Figure S8C, S8D**). Remarkably, spikes from all variants arising after BA.1 were substantially better at entering cells than WT, Beta, or Delta. When compared to WT, viral entry of Beta, Delta, BA.1, and BA.2 were all slightly enhanced, whereas variants from BA.5 to HK.3 infected 10-30 times more efficiently than WT (**Figure 6F**). Notably, BA.2.86 (which is derived from BA.2) was similar in its ability to enter cells as BA.2, but its descendant JN.1 has become the dominant circulating strain in the United States, with 15-fold greater efficiency at cell entry than WT. Taken together, these observations suggest that Bivalent and XBB.1.5 boosters substantially improve the neutralization activity of sera against highly mutated variants, but that there are substantial differences in the efficiency of spike mediated cell entry which has significantly increased in more recent strains.

## DISCUSSION

Although the COVID-19 pandemic declaration was ended by the WHO in May 2023, SARS-CoV-2 continues to transmit globally in the form of novel variants. We sought to understand how the constellation of mutations that have appeared since the beginning of the pandemic were impacted by the changing landscape of humoral immunity driven by updated vaccines. To this end, we determined the neutralization potency of patient sera against pseudoviruses representing each of the individual mutations observed within 50 strains of SARS-CoV-2 that have emerged, totaling 220 pseudoviruses. Our study utilized samples from 20-22 COVID naive donors who received a primary vaccination series, defined as either two shots of the Pfizer^18^ or Moderna^19^ mRNA vaccines, or a third mRNA booster dose. We also analyzed longitudinal samples from an older cohort consisting primarily of nursing home residents, taken after each successive vaccination. These individuals received up to a total of six mRNA vaccines, including the recent XBB.1.5 booster.

We find that sera from donors who received a primary vaccine series were especially vulnerable to individual mutations across multiple regions of the spike protein, including NTD, RBD, S1 and the C-terminus of S2. All individual mutations within the RBD resulted in escape from primary vaccination. Specifically, three receptor-binding domain (RBD) mutations from the Beta variant (K417N, E484K, N501Y) and two mutations from the Delta variant (L452R, P681R) substantially lowered the neutralization activity of sera from primary donors. Interestingly, sera from donors who received a third mRNA booster dose exhibited significantly improved neutralization across many, but not all, of the individual mutants we tested. New clusters of mutations formed compared to primary series clustering; notably, P681R and L452R clustered together and were among the hardest single mutants to neutralize, suggesting that these mutations were critical to the Delta variant overtaking other VOCs at the time. We observed that boosting disproportionately increased titers against specific key escape mutations across the spike including D80A, K417N, T572I and Q677H. Consistent with our previous report^13^, we found that boosting resulted in substantially greater breadth of neutralization against more recent strains. However, we observed a distinct loss of activity for boosted sera beginning with the emergence of the BQ.1.1 variant. Interestingly, this appeared to be largely driven by novel RBD mutations at positions 445 and 486, which have been retained in all variants since XBB, suggesting that they provide a significant advantage to the virus.

In response to the rapid emergence of SARS-CoV-2 variants, the vaccine formulation was twice updated; first to deliver a bivalent immunogen containing spike proteins from the WT and BA.4/BA.5 strain^50,51^ and then again to deliver a monovalent XBB.1.5 spike sequence ^48,52^. We find that the bivalent booster results in neutralizing activity comparable to wildtype for variants BA.5 and earlier, while also improving activity against BQ.1.1 and later variants, although these later variants still significantly escaped bivalent booster vaccination. In contrast, we find that the XBB.1.5 booster exhibits substantial improvement against later variants and only saw significant escape against the prior Beta variant and currently dominant JN.1 variant.

In addition to escape from vaccine sera, we found that mutations also contribute significantly to the ability of pseudovirus to infect cells, suggesting that variant selection is optimizing both antibody escape and viral entry. We found that spikes from variants post BA.1 produced pseudoviruses that were up to 30-fold better at transducing target cells than wild-type, suggesting that WT SARS-CoV-2 spike was not optimally configured for ACE2-dependent viral entry.

Our study demonstrates that individuals receiving the full mRNA vaccination regimen, including the XBB.1.5 booster, have maximally effective neutralizing activity against recent strains of SARS-CoV-2. However, despite improvements mediated by XBB.1.5 boosters against more recently emerging strains, there is still escape, particularly by the latest BA.2.86 descendant JN.1. This highlights the ongoing challenge presented by continued evolution of SARS-CoV-2, which has largely outpaced attempts to update vaccines. Our findings support the development of novel vaccine and prevention modalities capable of eliciting broad protection against future variants/outbreaks. Of note, a number of monoclonal antibodies have been described that target highly conserved epitopes such as the stem helix^53–56^ or fusion peptide^57–60^, which are capable of neutralizing across the entire Coronaviridae family. Next-generation vaccines that exploit these and other sites of vulnerability across Coronaviruses will likely be necessary to combat the ongoing evolution of SARS-CoV-2 and may help prevent future pandemics.

### Limitations of the study

While numerous prior studies have shown that neutralization activity against pseudoviruses is well-correlated to replication competent SARS-CoV-2^34–38^, it is possible that the mutations in the more recent variant spike proteins may cause them to behave differently than previously tested variants. In addition, while we confirmed that ACE2 expression is required for infection of 293T cells, natural target cells in the respiratory tract may express alternative receptors or attachment factors that facilitate infection and are not adequately modeled in our system. The study is limited to the evaluation of serum neutralizing antibodies to the spike protein for our pseudovirus neutralization assay, and does not take into account neutralizing antibodies to other proteins including the N and M proteins. Additionally, our infectivity studies only take into account the infectivity of spike mutations, not mutations in other proteins that contribute to viral infectivity. Furthermore, we did not assess other antibody-mediated functions such as complement deposition, antibody-dependent cellular cytotoxicity, or antibody-dependent cellular phagocytosis, which may contribute to protection even in the absence of neutralizing antibodies. We also did not assess the role of vaccine-elicited cellular immune responses mediated by T cells and NK cells, which are likely to play a key role in disease prevention for vaccine recipients.

## Data Availability

All data produced in the present study are available upon reasonable request to the authors

## ACKNOWLEDGEMENTS, FUNDING SUPPORT

We wish to thank Michael Farzan, PhD, for providing ACE2-expressing 293T cells. This work was supported by the Peter and Ann Lambertus Family Foundation. A.B.B. was supported by NIAID R01s AI174875, AI174276, the NIDA Avenir New Innovator Award DP2DA040254 a Massachusetts Consortium on Pathogenesis Readiness (MassCPR) grant and CDC subcontract 200-2016-91773-T.O.2.

## AUTHOR CONTRIBUTIONS

**Conceptualization:** A.R., Y.C., W.G.B., A.B.B.

**Methodology:** A.R., Y.C., K.S.D., M.L.S, E.L.

**Samples:** D.J.G., M.C.P., A.J.I., D.H.C., S.G., W.G.B.

**Investigation:** A.R., Y.C., C.J.L., K.S.D., M.L.S., E.L.

**Data Curation:** A.R., Y.C., K.S.D., M.L.S.

**Writing – Original Draft**: A.R.

**Writing – Reviewing and Editing:** A.R., Y.C., A.B.B. edited the paper with input from co-authors.

## INCLUSION AND DIVERSITY

We worked to ensure gender balance in the recruitment of human subjects. We worked to ensure ethnic or other types of diversity in the recruitment of human subjects. We worked to ensure that the study questionnaires were prepared in an inclusive way. One or more of the authors of this paper self-identifies as an underrepresented ethnic minority in science. One or more of the authors of this paper self-identifies as a member of the LGBTQ+ community.

## STAR * METHODS

### RESOURCE AVAILABILITY

#### Lead Contact

Further information and requests for resources and reagents should be directed to and will be fulfilled by Alejandro Balazs.

#### Materials Availability

Plasmids generated in this study will be available through Addgene. Recombinant proteins and antibodies are available from their respective sources.

#### Data and Code Availability

This study did not generate sequence data. Data generated in the current study (including neutralization measurements and flow cytometric files) have not been deposited in a public repository but are available from the corresponding author upon request. The code used to make the phylogenetic trees can be made available from the corresponding author upon request.

### EXPERIMENTAL MODEL AND SUBJECT DETAILS

#### Human subjects

Use of human samples was approved by Partners Institutional Review Board (protocol 2020P002274) and the WCG Institutional Review Board (protocol 1316159). Serum samples from 42 vaccine recipients that received two or three doses of the BNT162b2 or mRNA-1273 vaccine were collected. For each individual, basic demographic information including age and sex as well as any relevant COVID-19 history was obtained. The current analysis is part of an ongoing study ^61,62^ in which 29 NH residents and 7 healthcare workers (HCWs) are consented directly or through their legally authorized representative if needed and serially sampled before and after each SARS-CoV-2 vaccine dose. This analysis includes data collected between August 23, 2021, and September 8, 2022. HCWs worked at 3 NH buildings, 2 Cleveland area hospitals, and Case Western Reserve University. Residents and HCWs who received SARS-CoV-2 mRNA vaccines [(BNT162b2 (Pfizer-BioNTech) or mRNA-1273 (Moderna)] were included, and those who received the Ad26.COV2.S (Janssen) vaccine were excluded. Participants received their first booster dose 8–9 months after the primary vaccination series, and their second booster 4 to 6 months after the first booster. We report results from blood samples obtained at time points prior to and following the two booster doses.

#### Cell lines

HEK 293T cells (ATCC) were cultured in DMEM (Corning) containing 10% fetal bovine serum (VWR), and penicillin/streptomycin (Corning) at 37°C/5% CO_2_. 293T-ACE2 cells were a gift from Michael Farzan (Scripps Florida) and Nir Hacohen (Broad Institute) and were cultured under the same conditions. Confirmation of ACE2 expression in 293T-ACE2 cells was done via flow cytometry.

## METHOD DETAILS

### Construction of variant spike expression plasmids

To create variant spike expression plasmids, we performed multiple PCR fragment amplifications utilizing oligonucleotides containing each desired mutation (Integrated DNA Technology) and utilized overlapping fragment assembly to generate the full complement of mutations for each strain. Importantly we generate these mutations in the context of our previously described codon-optimized SARS-CoV-2 spike expression plasmid harboring a deletion of the C-terminal 18 amino acids that we previously demonstrated to result in higher pseudovirus titers. Assembled fragments were inserted into NotI/XbaI digested pTwist-CMV-BetaGlobin-WPRE-Neo vector utilizing the In-Fusion HD Cloning Kit (Takara). All resulting plasmid DNA utilized in the study was verified by whole-plasmid deep sequencing (Illumina or Primordium Labs) to confirm the presence of only the intended mutations.

### SARS-CoV-2 pseudovirus neutralization assay

To compare the neutralizing activity of vaccinee sera against coronaviruses, we produced lentiviral particles pseudotyped with different spike proteins as previously described (Garcia-Beltran et al. 2021). Briefly, pseudoviruses were produced in 293T cells by PEI transfection of a lentiviral backbone encoding CMV-Luciferase-IRES-ZsGreen as well as lentiviral helper plasmids and each spike variant expression plasmid. Following collection and filtering, production was quantified by titering via flow cytometry on 293T-ACE2 cells. Neutralization assays and readout were performed on a Fluent Automated Workstation (Tecan) liquid handler using 384-well plates (Grenier). Three-fold serial dilutions ranging from 1:12 to 1:8,748 were performed for each serum sample before adding 125–250 infectious units of pseudovirus for 1 h. Subsequently, 293T-ACE2 cells containing polybrene were added to each well and incubated at 37°C/5% CO_2_ for 48-60 hrs. Following transduction, cells were lysed using a luciferin-containing buffer (Siebring-van Olst et al. 2013) and shaken for 5 min prior to quantitation of luciferase expression within 1 h of buffer addition using a Spectramax L luminometer (Molecular Devices). Percent neutralization was determined by subtracting background luminescence measured in cell control wells (cells only) from sample wells and dividing by virus control wells (virus and cells only). Data was analyzed using Graphpad Prism and pNT_50_ values were calculated by taking the inverse of the 50% inhibitory concentration value for all samples with a neutralization value of 80% or higher at the highest concentration of serum.

### Titering

To determine the infectious units of pseudotyped lentiviral vectors, we plated 400,000 293T-ACE2 cells per well of a 12-well plate. 24 h later, three ten-fold serial dilutions of neat pseudovirus supernatant were made in 100 μL, which was then used to replace 100 μL of media on the plated cells. Cells were incubated for 48 h at 37°C/5% CO_2_ to allow for expression of ZsGreen reporter gene and harvested with Trypsin-EDTA (Corning). Cells were resuspended in PBS supplemented with 2% FBS (PBS+), and analyzed on a Stratedigm S1300Exi Flow Cytometer to determine the percentage of ZsGreen-expressing cells. Infectious units were calculated by determining the percentage of infected cells in wells exhibiting linear decreases in transduction and multiplying by the average number of cells per well determined at the initiation of the assay. At low MOI, each transduced ZsGreen cell was assumed to represent a single infectious unit.

### Quantitation of pseudovirus by RT-qPCR

To determine the genome copy concentration of pseudotyped lentiviral vectors, lentiviral RNA was extracted from pseudovirus supernatant using the QIAamp viral RNA mini kit (Qiagen). Each sample was serially diluted, and each dilution was treated with 1.2 U of Turbo DNase (Invitrogen) at 37°C for 30 min followed by heat inactivation at 75°C for 15 min. 10 μL of the treated RNA was used in a 20 μL qRT-PCR reaction with the qScript XLT one-step RT-qPCR Tough Mix, low ROX mix (Quanta Biosciences), a TaqMan probe containing locked nucleic acids (/56-FAM/AGC+C/i5NitInd/GG+GA/ZEN/GCTCTCTGGC/3IABkFQ/) (IDT), and primers designed for targeting the LTR gene of NL4-3 HIV genome, from which the lentiviral vector was derived (5′-GGTCTCTCTIGITAGACCAG and 3′-TTTATTGAGGCTTAAGCAGTGGG). Each dilution was run in duplicate on a QuantStudio 12K Flex (Applied Biosystems). The following cycling conditions were used: 50°C for 10 min, 95°C for 3 min followed by 50 cycles of 95°C for 15 s and 60°C for 1 min. Virus titer was determined by comparison with a standard curve generated using a plasmid standard generated from serial dilution of CMV-Luciferase-IRES-ZsGreen lentiviral backbone. DNase and No DNase controls were also included at 2.5 × 10^8^ GC/mL of the same plasmid. The range of the assay was from 2.5 × 10^7^ GC/mL to 1.5 × 10^3^ GC/mL. Upon analysis, the average of the three most concentrated dilutions within range of the standard were used to calculate genome copies/mL.

## QUANTIFICATION AND STATISTICAL ANALYSIS

Data and statistical analyses were performed using GraphPad Prism 9.0.1, and R v4.0.2. Flow cytometry data was analyzed using FlowJo 10.7.1. For all figures statistical significance was defined by GraphPad Prism as * = P<0.0332, ** = P<0.0021, ***= P<0.0002, ****= P<0.0001. Heatmaps and hierarchical clustering were performed using pheatmap package v1.0.12 in RStudio.

---

**Supplemental Figure 1:**
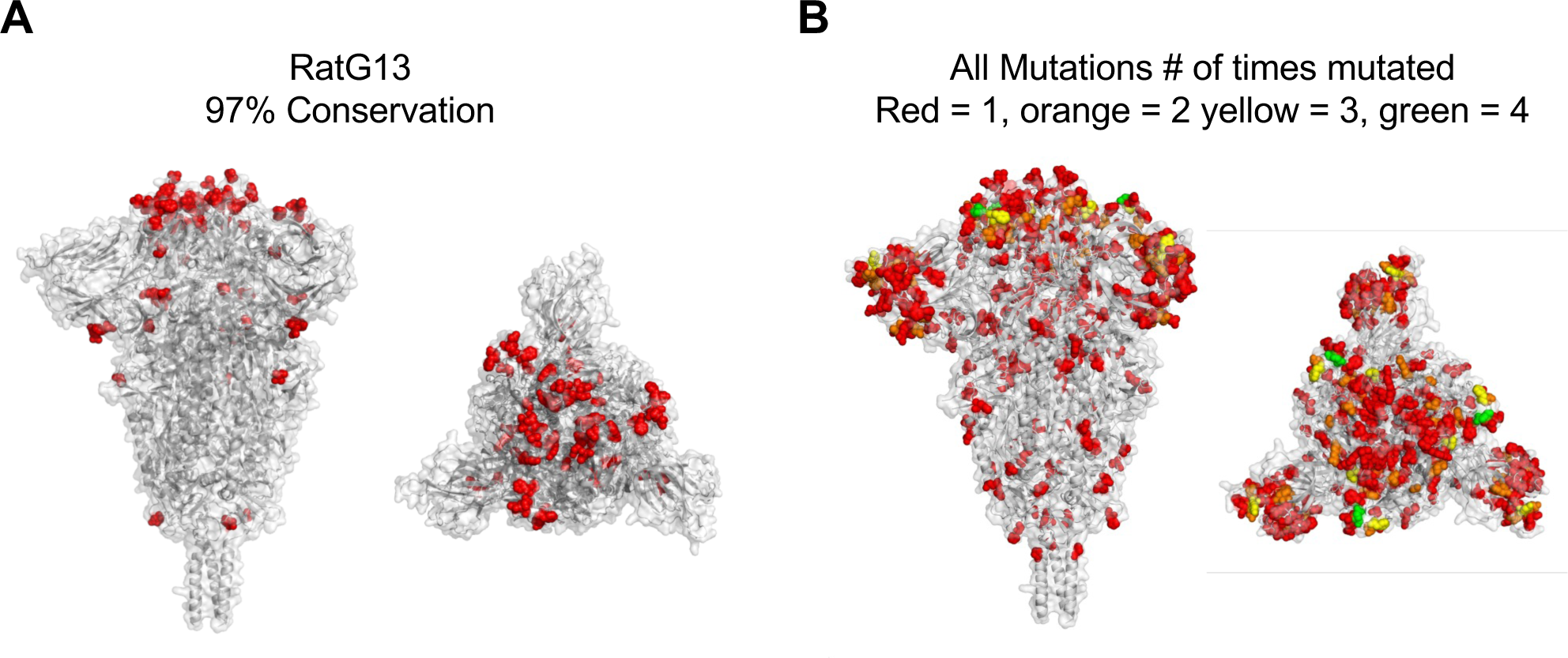
Variants of Concern have repeatedly mutated specific residues and have diversified beyond related pre-endemic coronaviruses. (A) Crystal structure of pre-fusion stabilized SARS-CoV-2 spike trimer (PDB ID 6XR8) with differences with the closely related RaTG13 coronavirus highlighted in red (Front and Top View). (B) Crystal structure of pre-fusion stabilized SARS-CoV-2 spike trimer (PDB ID 6XR8) with all 164 mutations found in variants of concern and variants of interest up-to and including JN.1, colored by the number of times they have been observed in a single variant of concern or variant of interest. Red = 1, orange = 2 yellow = 3, green = 4.

**Supplemental Figure 2:**
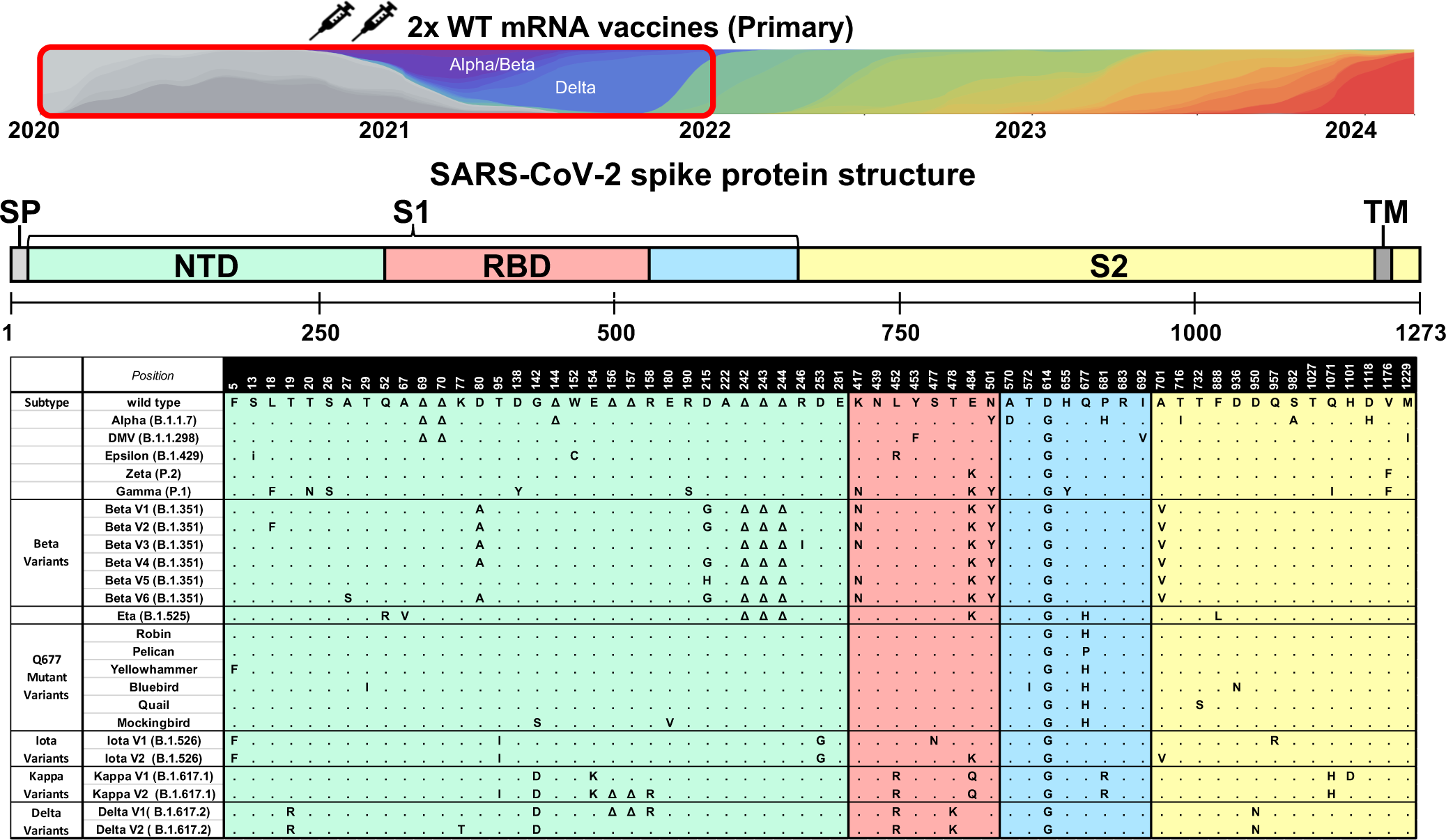
Mutations within variants through Delta tested against primary vaccinee sera. Schematic of SARS-CoV-2 spike protein with the mutations present in each of the variants tested in neutralization assays (**Fig 2)**. The mutations represent the consensus sequence for each strain. In the case of Beta (B.1.351), Iota (B.1.526), Kappa (B.1.617.1) and Delta (B.1.617.2), the 2-6 most abundant subvariants (V1-V6) deposited in GISAID were assessed. The following abbreviations are used: SP: signal peptide; NTD: N-terminal domain; RBD: receptor binding domain; TM: transmembrane domain.

**Supplemental Figure 3:**
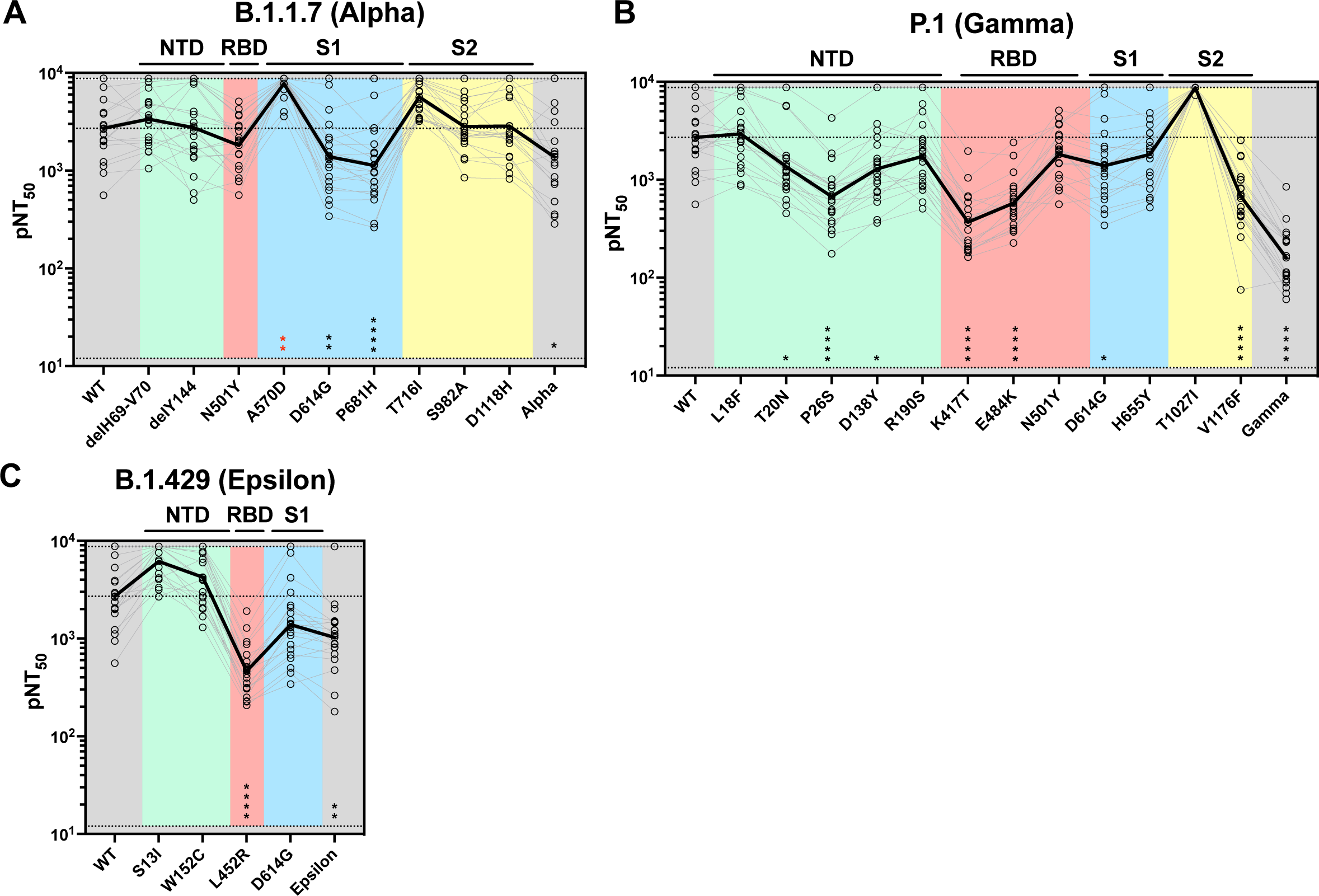
Individual mutations in VOC have modest impacts on and neutralization titers. (A-C) Pseudovirus neutralization titers for 24 primary vaccinee sera against individual mutant pseudoviruses and their respective strains harboring the combination of mutations Alpha (A), Gamma (B), and Epsilon (C). The solid black line represents geometric mean pNT50 for reference. Each pseudovirus was compared to wild type using a One Way ANOVA with Friedman’s Test and Dunn’s multiple comparisons test. (* = P<0.0332, ** = P<0.0021, ***= P<0.0002, ****= P<0.0001).

**Supplemental Figure 4:**
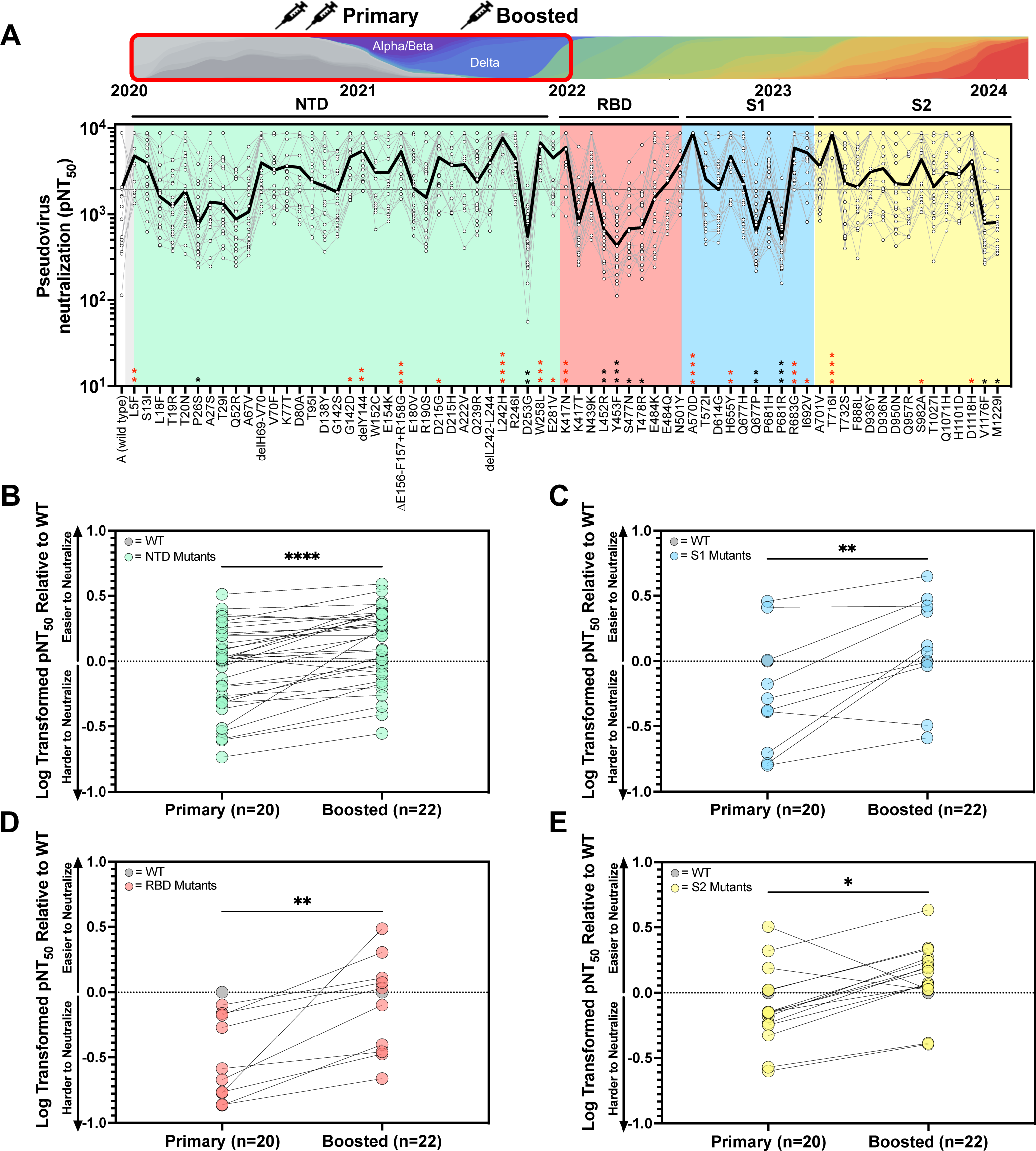
Booster vaccination yields humoral immunity capable of neutralizing most single mutants through the Delta variant of SARS-CoV-2. (A) (Top) Schematic illustrating the time period from which variants were selected and the vaccinee sera to be tested below. (Bottom) pNT_50_ are plotted for all individual spike mutations through Delta (1.617.2) for 24 COVID naive donors that received primary vaccination (2 doses of Moderna or Pfizer) and a booster shot. The solid black line represents geometric mean pNT_50_ for reference. The following abbreviations are used NTD = N-Terminal Domain, RBD = Receptor Binding Domain, S1 = S1 Subunit, S2 = S2 subunit. pNT_50_ of each spike mutant was compared to pNT_50_ of wild type using a One Way ANOVA with Friedman’s test and Dunn’s multiple comparisons test. (* = P<0.0332, ** = P<0.0021, ***= P<0.0002, ****= P<0.0001, red = significantly higher than WT). (B-E) Geometric mean pNT_50_ values for mutations separated by location on the spike protein normalized to wild-type and log transformed to compare primary to boosted values within: (B) NTD, (C) RBD, (D) S1 without NTD and (E) S2. Statistics represent Wilcoxon two-tailed paired, non-parametric t-test (* = P<0.0332, ** = P<0.0021, ***= P<0.0002, ****= P<0.0001).

**Supplemental Figure 5:**
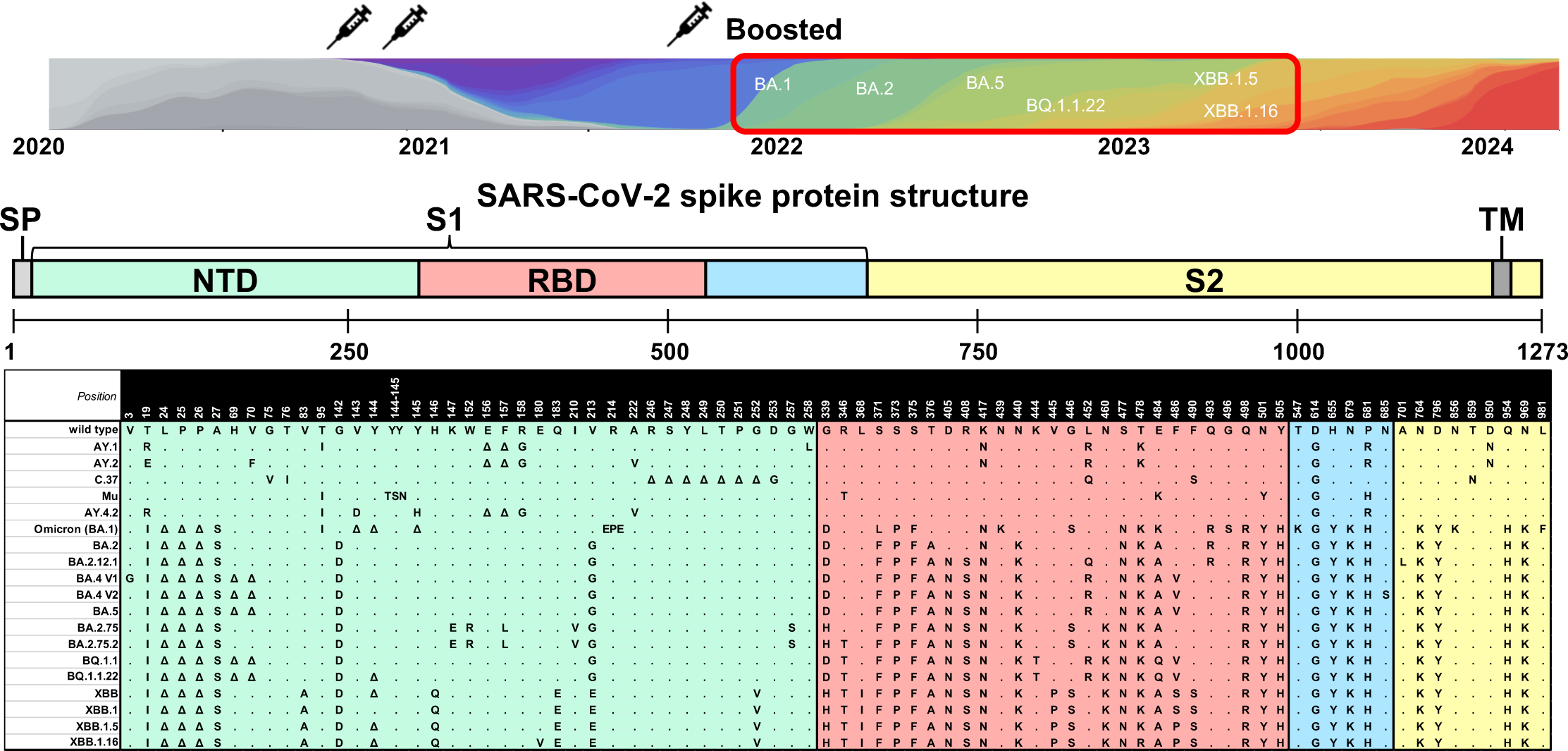
All mutations in strains post-Delta tested against boosted sera. All mutations observed in variants that emerged after Delta, up-to and including the XBB.1.16 variant.

**Supplemental Figure 6:**
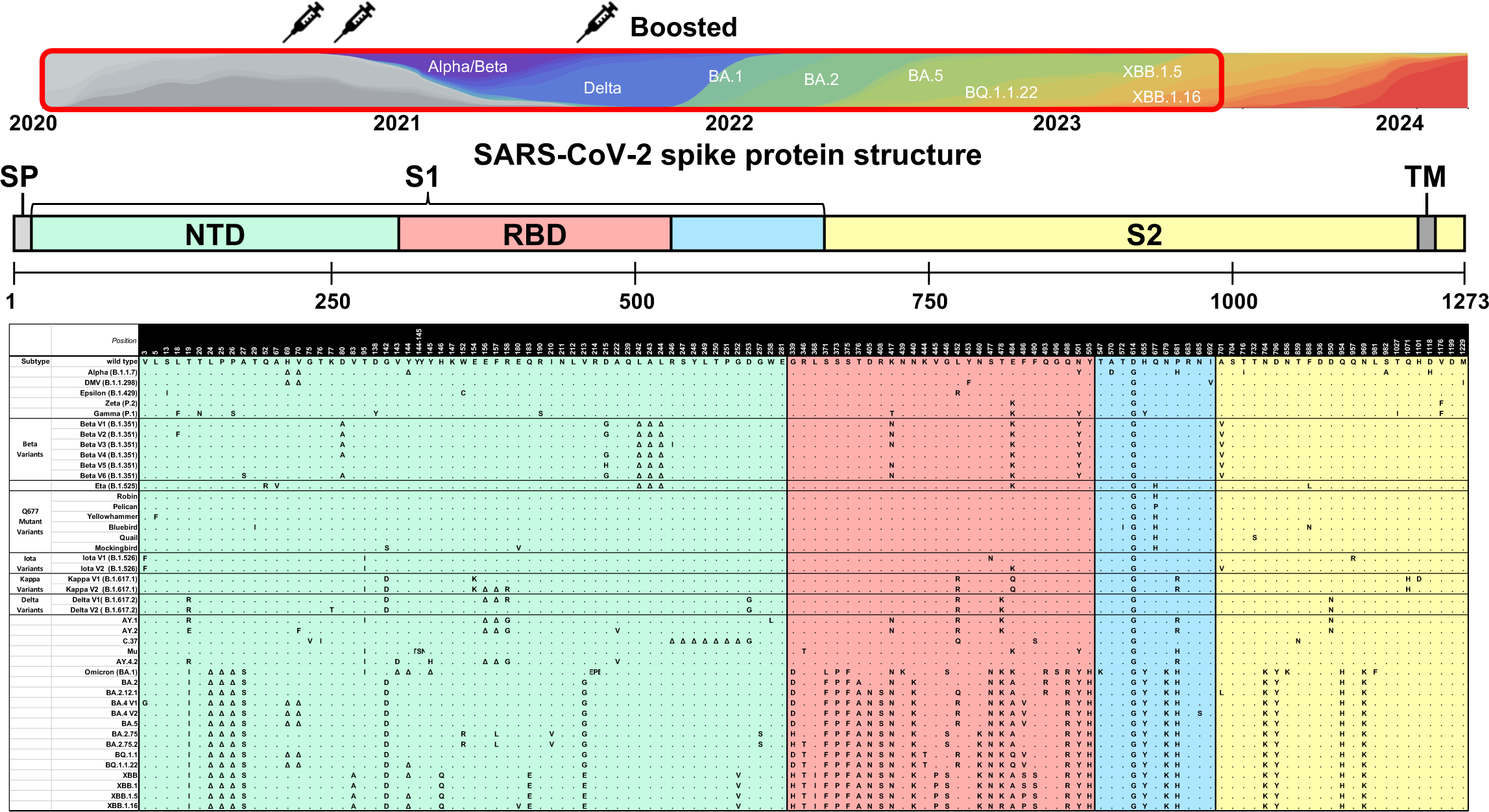
All mutations tested against boosted sera. All mutations and variants tested against 24 mRNA boosted donor samples.

**Supplemental Figure 7:**
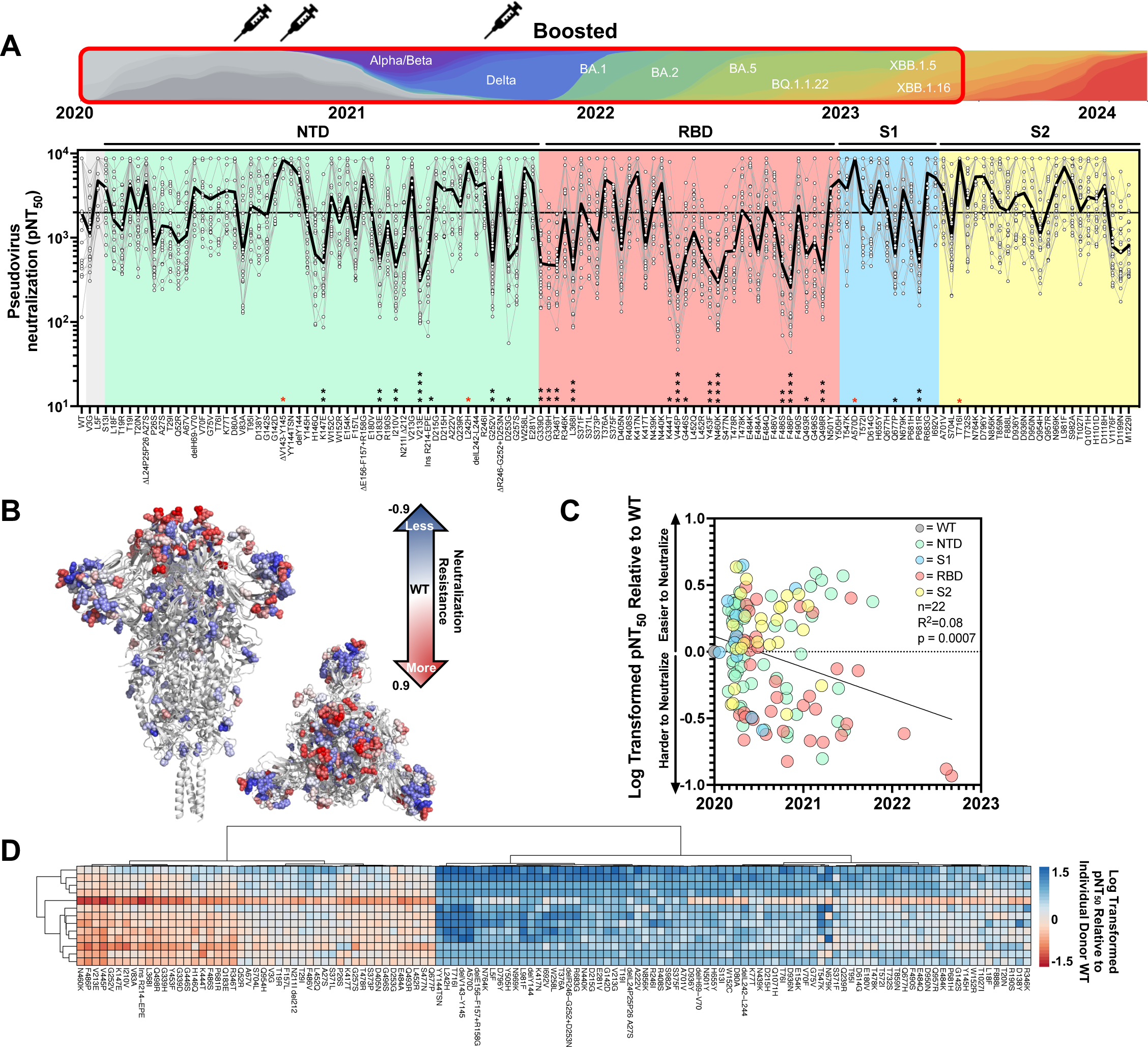
Novel spike mutations are able to significantly escape boosted donor sera. (A) (Top) Schematic illustrating the time period from which variants were selected and the vaccinee sera to be tested below. (Bottom) pNT_50_ are plotted for all individual spike mutations occurring from WT through XBB.1.16 for 24 COVID naive donors that received primary vaccination (2 doses of Moderna or Pfizer) and a booster shot. The solid black line indicates the geometric mean of the titers against each pseudovirus. The following abbreviations are used NTD = N-Terminal Domain, RBD = Receptor Binding Domain, S1 = S1 Subunit, S2 = S2 subunit. Each pseudovirus was compared to SARS-CoV-2 wild type using a One Way ANOVA with Kruskal Wallis Test and Dunn’s multiple comparisons test (* = P<0.0332, ** = P<0.0021, ***= P<0.0002, ****= P<0.0001). (B) The corresponding spike crystal structures (PDB 6xr8) from (A) with mutations colored according to neutralization resistance to boosted sera highlighting mutations between WT and XBB.1.16. Log transformed values for each mutation in the spike are plotted on a three color scale with a spectrum of -0.9 to 0.9. (C) Log transformed pNT_50_ geometric mean values relative to WT for boosted donor sera against all individual variants in strains through XBB.1.16 plotted against the date of first submission to GISAID and colored by spike region. (D) Two-way hierarchical clustering of pNT_50_ values of sera obtained from boosted donors (rows) across each variant (columns) relative to individual donor WT titer. pNT_50_ values are plotted in a heat map colored according to neutralizing activity relative to WT. Donors with WT neutralizing titers at the maximum of our assay were excluded. Clustering was performed using pheatmap package v1.0.12 in R-Studio.

**Supplemental Figure 8:**
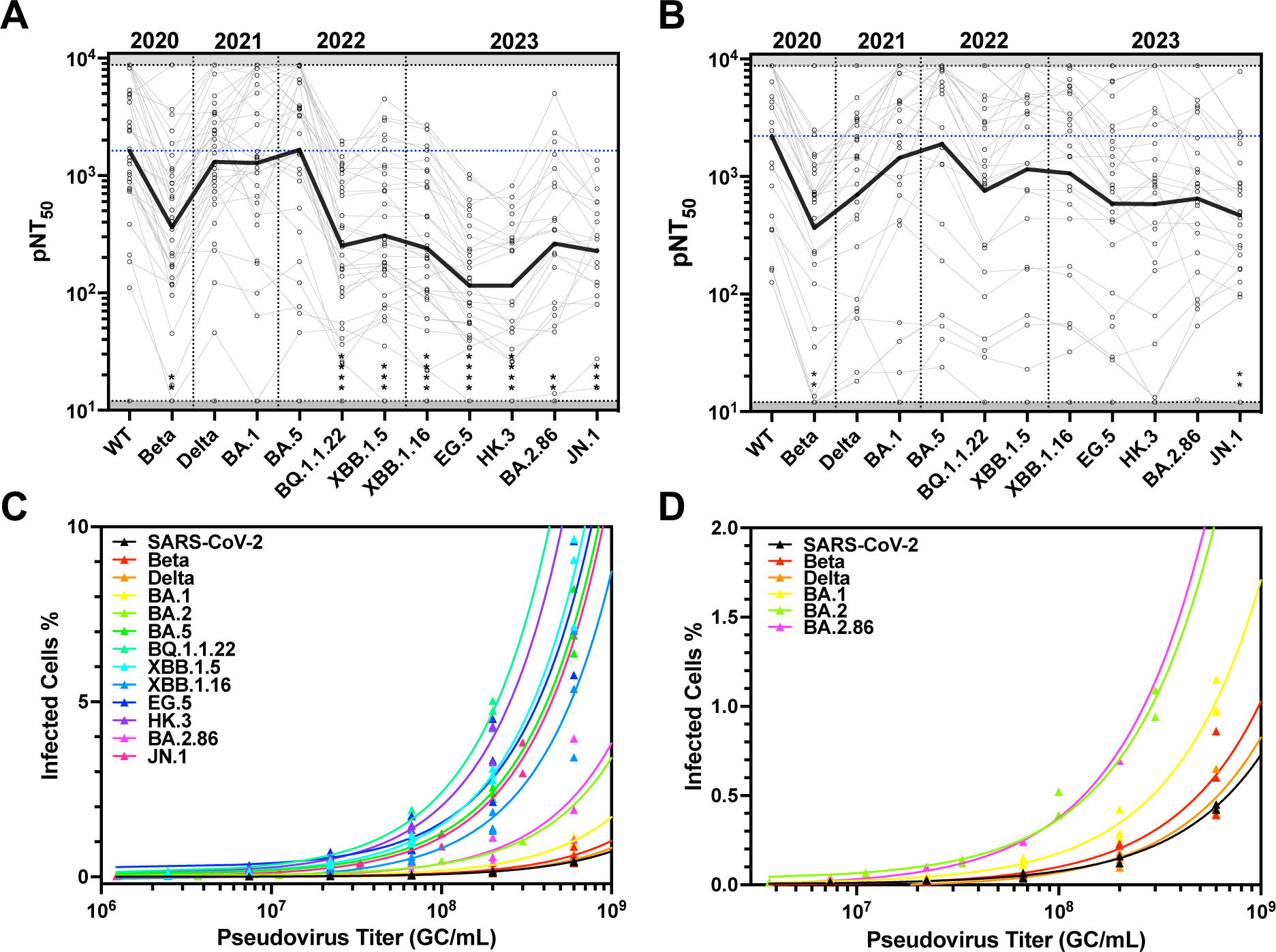
Bivalent boosters improve neutralization across pandemic variants, despite increasing infectivity. (A-B) pNT_50_ values of individual serum samples after (A) bivalent or (B) XBB.1.5 booster against each variant. Strains are plotted in the order in time that they first appear. The solid black line indicates the geometric mean of the titers against each pseudovirus. (C-D) Efficiency of viral entry of spike-pseudotyped lentiviruses into ACE2-293T cells across genome copies. Viruses were produced in duplicate to triplicate. qPCR titer was determined and used to normalize viruses to defined GC/mL values before being serially diluted and added to ACE2-293T target cells. Cultures exhibiting transduction above 10% or fewer than 5 transduced cells were excluded from analysis. (C) Includes all spikes tested while (D) shows a subset of viruses resulting in less than 1.5% transduction rates.

**Supplementary Table 1:** List of strains and numbers of mutations. All strains evaluated sorted by date of first appearance in GISAID with corresponding number of total spike mutations and RBD mutations denoted.

| Strain | First Submission to GISAID | # Spike Mutations | # RBD Mutations |
| --- | --- | --- | --- |
| Wuhan | 01-11-2020 | - | - |
| Robin | 04-06-2020 | 2 | 0 |
| Yellowhammer | 05-16-2020 | 3 | 0 |
| Pelican | 06-12-2020 | 2 | 0 |
| Alpha (B.1.1.7) | 10-14-2020 | 9 | 1 |
| DMV (B.1.1.298) | 11-05-2020 | 5 | 1 |
| Quail | 11-13-2020 | 3 | 0 |
| Epsilon (B.1.429) | 11-21-2020 | 4 | 1 |
| Beta (B.1.351 v1-v6) | 11-26-2020 | 7-9 | 2-3 |
| Zeta (P.2) | 12-02-2020 | 3 | 1 |
| Bluebird | 12-03-2020 | 5 | 0 |
| Iota (B.1.526) | 12-09-2020 | 6 | 1 |
| Eta (B.1.525) | 01-04-2021 | 7 | 1 |
| Mockingbird | 01-06-2021 | 4 | 0 |
| Gamma (P.1) | 01-10-2021 | 12 | 3 |
| Delta (B.1.617.2 v1-v2) | 01-30-2021 | 8 | 2 |
| C.37 | 02-10-2021 | 7 | 2 |
| Kappa (B.1.617.1 v1-v2) | 03-05-2021 | 8-9 | 2 |
| Mu (B.1.621) | 03-29-2021 | 7 | 3 |
| AY.1 | 05-03-2021 | 11 | 3 |
| AY.2 | 05-26-2021 | 11 | 3 |
| AY.4.2 | 06-11-2021 | 8 | 0 |
| Omicron (BA.1) | 11-22-2021 | 31 | 15 |
| BA.2 | 12-06-2021 | 27 | 14 |
| BA.2.12.1 | 02-10-2022 | 31 | 17 |
| BA.4 v1-v2 | 03-08-2022 | 32 | 17 |
| BA.5 | 03-15-2022 | 31 | 17 |
| BA.2.75 | 06-15-2022 | 35 | 17 |
| BQ.1.1 | 08-06-2022 | 34 | 20 |
| BA.2.75.2 | 09-05-2022 | 38 | 19 |
| BQ.1.1.22 | 09-11-2022 | 35 | 20 |
| XBB | 09-12-2022 | 40 | 22 |
| XBB.1 | 10-11-2022 | 39 | 22 |
| XBB.1.5 | 10-30-2022 | 40 | 22 |
| XBB.1.16 | 02-06-2023 | 41 | 22 |
| EG.5.1 | 03-28-2023 | 42 | 23 |
| HK.3 | 07-10-2023 | 43 | 24 |
| JD.1 | 07-11-2023 | 42 | 23 |
| BA.2.86 | 08-13-2023 | 57 | 26 |
| JN.1 | 09-20-2023 | 58 | 27 |

